# Evaluating the accessibility of exercise referral schemes to stroke survivors in England: A census of local authorities

**DOI:** 10.64898/2026.09.21.26363574

**Authors:** James Smith, Philip Nagy, David Tod, Carol Holland, Hannah Jarvis

**Affiliations:** Faculty of Health and Medicine, Lancaster University, Lancaster, United Kingdom

## Abstract

**Background:** Owing to a growing body of evidence, clinical guidelines now recommend exercise for stroke survivors. However, many stroke survivors are physically inactive and may need more support to engage with exercise in the community. There are between 600-800 exercise referral schemes in the United Kingdom, but engagement from stroke survivors is very low. This study aimed to evaluate the accessibility of exercise referral schemes in England to stroke survivors.

**Methods:** A cross-sectional, online survey was conducted throughout England. All 317 local authorities in England were invited to participate. Geospatial mapping was used to visualise the accessibility of exercise referral schemes across England. Data on scheme design and delivery parameters were analysed descriptively.

**Results:** Data were collected from 147 local authorities. Exercise referral schemes were offered in 134 local authorities, of which, stroke survivors were able to access 128. However, stroke-specific schemes were available in only 17 local authorities. Significant variations in exercise referral scheme design and delivery were identified across the country. Only two consistencies were found between schemes: the involvement of fitness instructors, and the lack of transport provision. The capacity of exercise referral schemes did not correlate with the general population, stroke population, or stroke prevalence within a local authority.

**Conclusions:** Exercise referral schemes are widely available across England, and the majority are open to stroke survivors. However, stroke-specific services are very limited and there is substantial variation in scheme eligibility, capacity, costs, and referral pathways. These inconsistencies, paired with the lack of transport provision, may explain the low uptake of exercise referral schemes within the stroke population.

## Background

There are over 1.5 million people living with the effects of stroke in the United Kingdom (UK),^1^ which is expected to grow to 2,119,400 people by 2035.^2^ Evidence of this growing burden has already been observed in England, with a 24% increase in stroke hospital admissions from 2004 to 2024.^3^ Stroke can cause both physical and psychological impairments, which can both be effectively rehabilitated using exercise.^4–6^ Owing to a growing body of evidence, national clinical guidelines now recommend exercise for stroke survivors.^7,8^ Despite these recommendations, many stroke survivors are physically inactive,^9^ and may need more support to engage with exercise in the community.

Exercise referral schemes (ERS) were first established in the UK in the 1990s as a public health intervention for the general population.^10^ Formal guidance,^11^ and specific clinical guidelines have been published,^12^ but significant variation in the delivery of ERS has been identified across England and the UK.^13,14^ The National Institute of Health and Care Excellence (NICE) guideline PH54 states that ERS should only be funded for people who are physically inactive and have an existing health condition,^12^ making many stroke survivors eligible for these schemes. Very little stroke specific research has been conducted in the context of ERS, with only two studies previously published in this area.^15,16^ Both studies explored experiences of stroke stakeholders involved with an exercise referral scheme. All stakeholders reported benefits to stroke survivors, such as improved physical function, independence and confidence.^15,16^ However, some safety concerns were raised.^15^

The uptake of stroke survivors in ERS schemes is extremely low, despite the established benefits and guideline recommendations. The National ReferAll Database contains data from 39,283 unique participants of ERS in the UK.^17^ Only 54 (0.1%) service users within the ReferAll Database participated in a stroke rehab programme, suggesting exercise referral schemes are a underutilised resource for stroke survivors. Best et al. investigated community exercise programmes for stroke survivors across Scotland, and found only four stroke-specific local authority run programmes.^18^ The total number of ERS in the UK is currently unknown; however, a recent study identified 625 schemes, demonstrating extensive provision.^14^ These schemes are an underutilised resource in the UK stroke population, but the reasons for this are unclear. The accessibility of ERS to stroke survivors is currently unknown and may be a key factor driving poor engagement from stroke survivors.

The primary aim of this study was to evaluate the accessibility of ERS across England to stroke survivors. Secondary aims were to: map current exercise referral schemes based on geographical location, compare delivery approaches, and evaluate service capacity.

## Methods

### Design

A cross-sectional online survey was conducted from 14/08/2025 to 02/03/2026. Ethical approval was granted by the Lancaster University Faculty of Health and Medicine Research Ethics Committee (FHM-2024-4432-RECR-2). All participants provided informed consent. The study was conducted in accordance with the Declaration of Helsinki.

### Participants

All 317 local authorities in England were invited to participate in the study.^19^ Given differing local government structures in Scotland, Wales, and Northern Ireland these areas were not included. A similar study has already been conducted in Scotland.^18^ Local authorities in England can either be two tier (county councils and district councils) or single tier (unitary authorities, metropolitan districts, London boroughs, City of London, and Isles of Scilly).^19^ In two tier authorities, council services are shared between the county council and its constituent district councils, whereas single tier authorities are responsible for all council services.^20^ The distribution of responsibilities between county councils and district councils is not standardised, so both council types were included in the study. If a county council provided a response, it also represented its constituent district councils.

### Recruitment

A census approach was taken, in which the whole population of local authorities were sampled.^21^ Contact details for all 317 local authorities were extracted from the specific local authorities website, and recruitment materials and links to the online survey were then sent via email. Some local authorities did not provide contact details but instead had an online contact form. In these instances, the recruitment information and link to the survey were sent via the online form. After the initial invitation, a reminder email was sent after approximately six weeks. If no response was recorded after a further six weeks from the reminder, an alternative contact was sought (i.e., a different team within the council). The process was then repeated with the alternative contact.

### Survey development

The online survey was developed specifically for this study in accordance with Dillman’s Tailored Design Method,^22^ using Qualtrics XM (Qualtrics, 2024). Previous surveys in this area were used to inform design.^13,18^ The survey was piloted with staff from a local authority owned leisure centre before recruitment. A consent form was integrated into the survey, which had to be completed before entering the survey proper. The survey was broken down into four sections. Section one identified the local authority responding then established if they ran an ERS (or similar programme) and if stroke survivors were able to access this scheme. If the respondent answered no to either question, the survey was ended. Section two concerned general scheme details, such as intended population, funding, partner organisations, and facilities. Section three concerned stroke-specific information, such as eligibility criteria, referral mechanisms, assessment, instructor qualifications, and cost to service users. Section four identified ERS delivery parameters, such as duration, services available, service capacity, session frequency, session duration, educational components, and transport provision. A complete HTML version of the survey is available in the supplementary materials.

### Data analysis

Data were exported from Qualtrics into Microsoft Excel, where data cleaning was conducted. Written responses were coded using content analysis by JS. Data were then imported into RStudio (version 4.4.1, 2024) for analysis. For nominal and ordinal data, frequencies and proportions were reported, and for continuous data, mean and standard deviation.

Geospatial mapping was used to visually display the accessibility of exercise referral schemes across England. Local authority boundaries were obtained from the Office for National Statistics Open Geography Portal.^23^ Accessibility was plotted based on five categories: no data, no ERS, ERS not open to stroke survivors, ERS open to stroke survivors, and stroke-specific ERS. A choropleth map was produced in RStudio (version 4.4.1, 2024) using the ‘sf’, ‘ggplot2’, ‘dplyr’, and ‘patchwork’ packages.

To assess if the capacity of ERS reflected need within the local authority, Spearman’s rho was used to correlate data from the British Heart Foundation Heart and Circulatory Disease Statistics 2025 to the capacity data captured from this survey.^24^ Nine individual analyses were conducted: general population and number of venues, general population and capacity of 1-2-1 sessions, general population and capacity of group sessions, stroke population and number of venues, stroke population and capacity of 1-2-1 sessions, stroke population and capacity of group sessions, stroke prevalence and number of venues, stroke prevalence and capacity of 1-2-1 sessions, and stroke prevalence and capacity of group sessions. An alpha of 0.05 was used and rho (ρ) was interpreted as: 0.00-0.29 – poor, 0.30-0.59 – fair, 0.60-0.79 – moderately strong, and >0.8 – very strong.^25^

## Results

Data were collected from 147 (46%) of the 317 local authorities across England. The frequencies and proportions for all variables are presented in Table 1. When stratified by region, response rate ranged from 59% (South East) to 25% (North East). Of the 147 responding local authorities, 134 reported offering an ERS. Stroke survivors were able to access ERS in 128 of these local authorities; however, stroke-specific ERS were only available across 17 local authorities (Figure 1). In six local authorities (South Norfolk, Manchester, Dudley, Crawley, Broadland, and Amber Valley), two separate ERS were available, resulting in a total of 133 schemes, across 127 local authorities, accessible to stroke survivors. In areas with two-tier council structures, the responsibility of providing ERS was not consistent with county councils or district councils. Hampshire, Hertfordshire, Leicestershire, Surrey, and Warwickshire County Councils reported providing ERS at a county level, whereas Devon, and Kent County Council did not. Crawley (non-metropolitan district council) reported receiving funding from West Sussex County Council, but it was unclear if this funding was provided to all districts in the county. The remaining 13 county councils did not respond.

**Figure 1.**
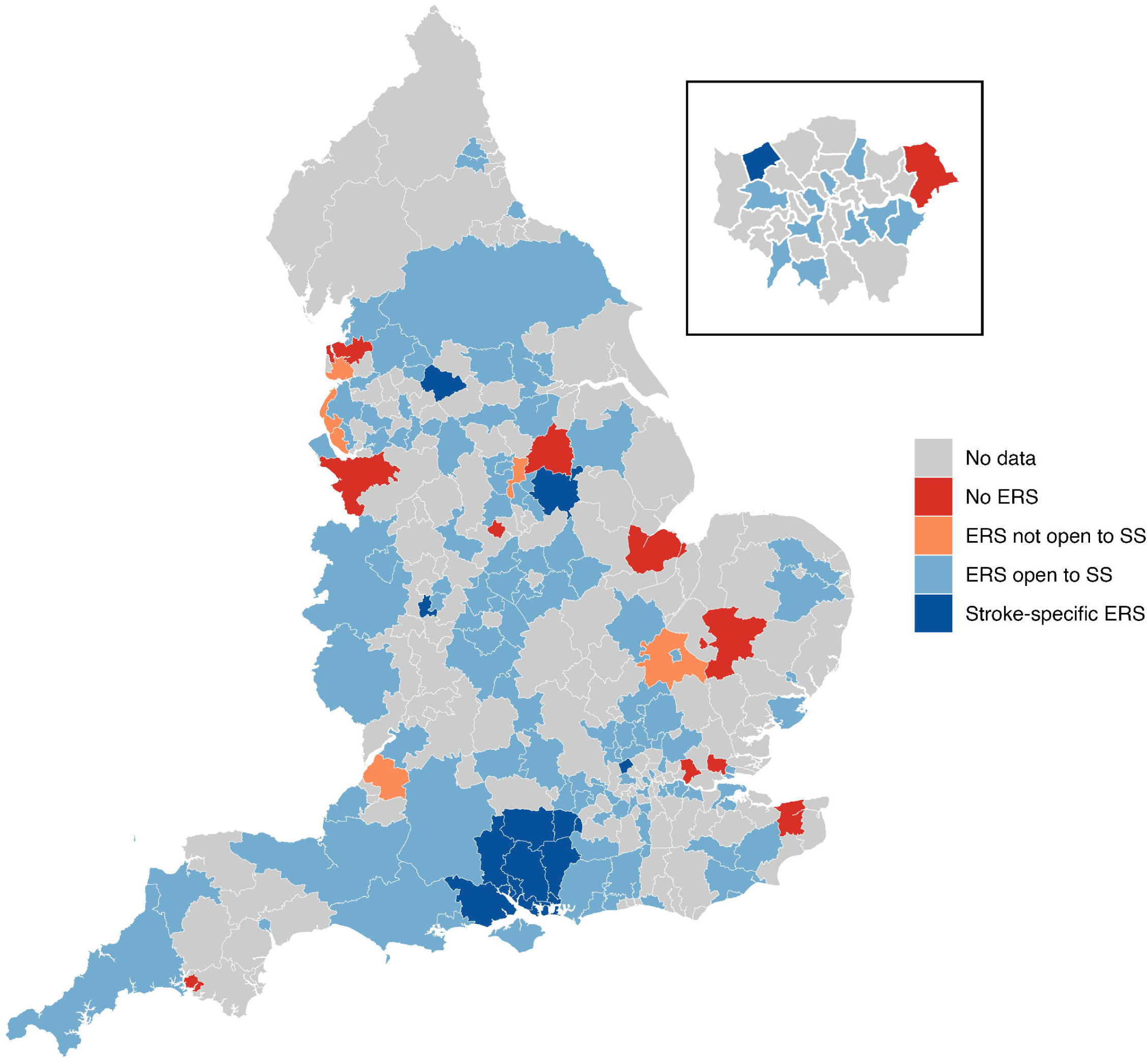
Availability of Exercise Referral Schemes to stroke survivors in England

**Table 1.** Descriptive statistics of survey responses.

| Domain |  |  | Frequency (%) |
| --- | --- | --- | --- |
| Stroke accessibility |  |  |  |
|  | Complete responses |  | 147 |
|  |  | East Midlands | 22 (56) |
|  |  | East of England | 22 (45) |
|  |  | London | 12 (36) |
|  |  | North East | 3 (25) |
|  |  | North West | 19 (53) |
|  |  | South East | 41 (59) |
|  |  | South West | 12 (40) |
|  |  | West Midlands | 11 (33) |
|  |  | Yorkshire and Humber | 5 (33) |
| Funding sources |  |  |  |
|  | Complete responses |  | 102 |
|  |  | Local authority | 62 (61) |
|  |  | Fees | 62 (61) |
|  |  | NHS | 25 (25) |
|  |  | Public health | 9 (9) |
|  |  | Leisure providers | 7 (7) |
|  |  | Charities | 6 (6) |
|  |  | Private organisations | 6 (6) |
|  |  | Sport England | 1 (1) |
|  |  | National Lottery | 1 (1) |
| Number of venues |  |  |  |
|  | Complete responses |  | 100 |
|  |  | 1-2 | 28 (28) |
|  |  | 3-4 | 27 (27) |
|  |  | 5-6 | 15 (15) |
|  |  | 7-8 | 5 (5) |
|  |  | 9-10 | 5 (5) |
|  |  | 11+ | 20 (20) |
| Number of sessions per week |  |  |  |
|  | Complete responses |  | 91 |
|  |  | >1 | 2 (2) |
|  |  | 1 | 10 (11) |
|  |  | 2 | 17 (19) |
|  |  | 3 | 7 (8) |
|  |  | 4 | 6 (7) |
|  |  | 5 | 6 (7) |
|  |  | 6 | 4 (4) |
|  |  | 7 | 0 (0) |
|  |  | >7 | 39 (43) |
| 1-2-1 service capacity |  |  |  |
|  | Complete responses |  | 32 |
|  |  | 1-10 | 20 (63) |
|  |  | 11-20 | 3 (9) |
|  |  | 21-30 | 2 (6) |
|  |  | 31-40 | 1 (1) |
|  |  | 41-50 | 2 (1) |
|  |  | 51+ | 4 (1) |
| Group service capacity |  |  |  |
|  | Complete responses |  | 85 |
|  |  | 1-25 | 38 (45) |
|  |  | 26-50 | 13 (15) |
|  |  | 51-75 | 6 (7) |
|  |  | 76-100 | 5 (6) |
|  |  | 101+ | 23 (27) |
| Referral sources |  |  |  |
|  | Complete responses |  | 98 |
|  |  | Health professional only | 54 (55) |
|  |  | Health professional or self-referral | 42 (43) |
|  |  | Self-referral only | 2 (2) |
| Transport provision |  |  |  |
|  | Complete responses |  | 94 |
|  |  | Provided | 3 (3) |
|  |  | Not provided | 91 (97) |
| Staffing |  |  |  |
|  | Complete responses |  | 98 |
|  |  | Fitness instructors | 96 (98) |
|  |  | Exercise physiologists | 12 (12) |
|  |  | Physiotherapists | 5 (5) |
|  |  | Occupational therapists | 2 (2) |
|  |  | Therapy assistants | 3 (3) |
|  |  | Nurses | 1 (1) |
|  |  | Swim instructors | 1 (1) |
|  |  | Sports coaches | 1 (1) |
|  |  | Wellbeing practitioner | 1 (1) |
|  |  | Volunteers | 1 (1) |
| Staff to service user ratio |  |  |  |
|  | Complete responses |  | 87 |
|  |  | 1:1-2 | 8 (9) |
|  |  | 1:3-4 | 3 (3) |
|  |  | 1:5-6 | 5 (6) |
|  |  | 1:7-8 | 7 (8) |
|  |  | 1:9-10 | 21 (24) |
|  |  | 1:11+ | 43 (49) |
| Programme duration |  |  |  |
|  | Complete responses |  | 97 |
|  |  | 5-8 weeks | 2 (2) |
|  |  | 9-12 weeks | 56 (58) |
|  |  | 13-16 weeks | 5 (5) |
|  |  | 17+ weeks | 34 (35) |
| Session duration |  |  |  |
|  | Complete responses |  | 91 |
|  |  | 16-30 minutes | 2 (2) |
|  |  | 31-45 minutes | 16 (18) |
|  |  | 46-60 minutes | 72 (79) |
|  |  | 61+ minutes | 1 (1) |
| Education |  |  |  |
|  | Complete responses |  | 94 |
|  |  | Provided | 53 (56) |
|  |  | Not provided | 41 (44) |

## Funding

The funding of community stroke exercise services was highly varied across ERS. Sources of funding were reported in 102 responses. The two most common sources of funding were from the local authorities (n=62), and fees paid by service users (n=62). Other sources of funding were: the National Health Service (n=25), public health organisations (n=9), not-for-profit leisure providers (n=7), charities (n=6), private organisations (n=6), Sport England (n=1), and the National Lottery (n=1). It was common for ERS to have multiple sources of funding (n=55). However, 27 ERS were funded solely by fees paid by service users, with no subsidisation from other sources.

Payment structures were reported in 99 responses. Service users were able to access 27 of the reported ERS free of charge, and 72 required payments. For the paid ERS, a variety of payment structures were reported, which included: complete upfront payment for the whole scheme, pay as you go, and monthly direct debit. Some ERS also required service users to pay a consultation/ joining fee before enrolling. Monthly prices were reported for 28 ERS. Costs to service users ranged from £15 to £38.50 per month, with a mean (standard deviation) of £27.67 (5.07). Pay as you go prices were reported by 34 ERS, and ranged from £2 to £7 per session, with a mean (standard deviation) of £4.47 (1.04). Complete upfront payment costs were reported by nine ERS. Price ranged from £12 to £97.50 for a 12-week scheme, with a mean (standard deviation) of £55.82 (29.29). Most ERS offer a choice of payment options to service users. Prices also varied within a given local authority, depending on the venue. Given the heterogeneity of pricing structures, and low number of responses in certain regions, it was not possible to compare prices across regions of England.

### Service capacity

The number of venues providing exercise services were reported by 100 ERS. There was a poor correlation between the number of venues and general population in a local authority (ρ=0.26; p=0.01), as well as population of stroke survivors (ρ=0.29; p<0.01). A fair correlation was found with stroke prevalence, but this was not statistically significant (ρ=0.32; p=0.76). The venues used by these schemes included: local authority owned leisure centres, privately owned leisure centres, leisure centres owned by charitable trusts, sports clubs, public outdoor spaces, community centres, sports pavilions, church halls, rehabilitation hospitals, privately owned rehabilitation facilities, care homes, and GP surgeries.

The number of exercise sessions (either group-based or one-to-one) that can be accessed by service users per week was reported by 91 ERS. The most common response was eight or more sessions per week (n=39), followed by two sessions per week (n=17).

One-to-one training was available to service users in 36 of the ERS, of which 32 reported the weekly capacity for one-to-one sessions. Most of these schemes (n=20) had capacity for 1-10 service users per week. However, others had capacity for more than 50 service users per week. No significant correlation was found with general population (ρ=0.29; p=0.11), or stroke population (ρ=0.10; p=0.60). However, a fair inverse correlation was found with stroke prevalence (ρ=-0.39; p=0.02).

Group exercise sessions were available in 94 of the reported ERS, of which 85 reported service capacity for group sessions. The two most common responses were at both ends of the scale: 1-25 (n=38) and >100 (n=23). No significant correlation was found with general population (ρ=0.03; p=0.78), stroke population (ρ=0.01; p=0.92), or stroke prevalence (ρ=-0.06; p=0.55).

### Accessing schemes

Referral mechanisms were reported by 98 ERS. Reported referral sources included health professionals, self-referrals, charities, and public health teams. For ERS that accept referrals from health professionals, only eight required specific professionals to make the referral. Four of these only allowed referrals from a general practitioner, three from a general practitioner or physiotherapist, and one from physiotherapists or occupational therapists. Some ERS also allowed referrals from non-healthcare professionals, such as, social prescribers, health and social care workers, fitness instructors, health and wellbeing coaches, and community workers.

Eligibility criteria were reported by 99 ERS. Eight ERS were reported to be open to anyone, eleven open to anyone who is physically inactive, and nine open to people living with a long-term health condition. In five schemes, eligible participants must have been at high risk of falls. Eligibility related to stroke chronicity was reported in 18 ERS; however, these criteria were not consistent. In ten schemes, a stroke must have occurred greater than three months ago, in one greater than six months, and another greater than twelve months. For an upper time limit, five schemes reported that a stroke must have occurred less than 12 months ago, and another reported less than 24 months ago. Other eligibility criteria included: being medically stable (n=33), able to follow instructions (n=5), and completed a phase three rehabilitation programme (n=2). Five ERS did not report specific criteria and instead conducted case-by-case eligibility assessments. Commonly reported exclusion criteria were: contraindications to exercise (n=10), impaired cognitive function (n=9), and wheelchair users (n=4).

Transport provision was reported by 94 ERS. The vast majority (n=91) did not provide any support with transport for service users. One scheme reported providing transport for service users, and a further two schemes reported providing transport depending on the individuals’ circumstances (i.e., where they live, level of physical function, financial circumstances).

### Staffing

Of the 98 ERS that reported staffing, fitness instructors were involved with 96. The minimum required qualification for a fitness instructor to be involved with the ERS was reported in 79 responses. A level three qualification was required in 52 schemes, and a level four qualification in 27. Only 17 ERS reported the use of a multidisciplinary team, with 79 being run exclusively by fitness instructors and 2 by exercise physiologists. The ratio of staff to participants was reported by 87 schemes, with most having ratios greater than 1:9 (n=64).

### Exercise parameters

Service users were able to access a wide range of exercise services as part of ERS, which included: gym-based exercise, water-based exercise, cycling, walking, team sports, racket sports, yoga, Pilates, and tai-chi. The length of ERS ranged from five to over sixteen weeks. The length of each exercise session was most commonly between 46-60 minutes (n=72). However, two schemes reported a session length of 16-30 minutes, 16 reported 31-45 minutes, and 1 reported session length greater than 60 minutes. Educational components were included in 53 ERS but not included in 41. Topics of education were highly varied across schemes, and included: physical activity, nutrition, weight management, behaviour change, smoking cessation, alcohol consumption, mental health, loneliness, pain, falls, goal setting, finances, housing issues, and raising awareness of other local health services.

## Discussion

This is the first study to evaluate the accessibility of ERS in England to stroke survivors and has a comparable sample size to a previous audit of general ERS published by the British Heart Foundation.^26^ The vast majority of responding local authorities offered an ERS that is open to stroke survivors; however, stroke-specific ERS were rare. Substantial variation in ERS funding, eligibility, referral pathways, staffing, and delivery were identified across the country. Ony two consistencies were found across ERS: the involvement of fitness instructors, and the lack of transport provision. The capacity of ERS did not reflect the need of either the general or stroke population within the local authority.

The vast majority of responding local authorities run ERS that are open to stroke survivors, but these services may not necessarily be accessible to stroke survivors. Despite stroke survivors not being explicitly excluded, the sequalae of stroke will result in many stroke survivors being ineligible for ERS. Exclusion criteria such as wheelchair users and cognitive impairments will disproportionately affect stroke survivors more than other populations.^27,28^ Furthermore, the delivery parameters employed by ERS in England, such as high numbers of service users to supervisors are inappropriate for all but the most mildly affected stroke survivors. The same findings have also been reported in Scotland.^18^ Stroke-specific exercise guidelines recommend a maximum ratio of six patients to one supervisor, due to the greater needs of the population.^29^ The lack of transport provision is another barrier that will prevent many stroke survivors accessing ERS.^30^ Despite being open to stroke survivors, most ERS in England are not accessible to stroke survivors.

Another factor that may be limiting accessibility for stroke survivors is cost. The cost to participate in an ERS was highly varied across the country, costing up to £38.50 per month (£115.50 for a 12-week programme). Almost half of UK stroke survivors experience financial hardship as a result of their stroke,^31^ and the price of community exercise services has been identified as the main barrier to stroke survivors exercising.^30^ The high subscription costs could be perpetuating health inequalities, as stroke survivors of lower socioeconomic status are less likely to access ERS. Given the established clinical benefits of exercise, in both the acute and chronic stages of stroke,^4–6^ this reduced access may influence clinical outcomes. Although there is no research specific to community exercise services, lower socioeconomic status is associated with reduced access to healthcare.^32^ In England, offering free access to ERS is feasible, as 27% of respondents are already doing this. There is a need to standardise free access to ERS in England to mitigate health inequalities that may be being propagated by current practices.

A small number of local authorities run stroke-specific ERS, which are specifically designed for the needs of stroke survivors. Although these are an excellent resource for stroke survivors,^16^ their inconsistent distribution across the country is causing inequitable access. A similar study in Scotland also found a small number of stroke-specific exercise programmes across the country.^18^ The Stroke Association have acknowledged that access to community services is a ‘postcode lottery’,^33^ and the findings of this study support this. As Figure 1 shows, there are areas with excellent provision for stroke survivors, such as Hampshire, whereas other areas have no ERS at all. Therefore, stroke survivors living in areas with better service provision have more opportunity to achieve better outcomes. Data from the ReferAll database shows that stroke-specific ERS have been running since at least 2012,^17^ and the fact that these services still exist demonstrates their sustainability. Given the recent changes to stroke management guidelines, stroke survivors living in areas with poor ERS provision may not have access to services recommended by The National Clinical Guideline for Stroke.^7^ Expansion of stroke-specific community exercise service is needed to meet current clinical guidelines and to combat the ‘postcode lottery’ access to rehabilitation.

The capacity of ERS across England were highly varied. Although the number of venues offering services within a scheme weakly correlated to both the general population and population of stroke survivors within a local authority, no correlation was found with either one-to-one or group capacity. However, the interpretation of one-to-one capacity should be interpreted with caution due to the low number of responses. These findings suggest that service capacity is not aligned to the need of the local population. Instead, capacity may be dictated by other factors such as previous funding, availability of facilities, priorities of the local authority, or the involvement of third-party leisure contractors. Given the growing population of stroke survivors in the UK,^2^ the capacity of community services are becoming increasingly important. A needs-based approach to commissioning ensures that areas with high stroke prevalence can accommodate more stroke survivors to ensure more equitable access to services across the country.

Although current ERS clinical guidelines do not provide recommendations for the content or structure of exercise, community based exercise guidelines for stroke are available.^34^ These guidelines suggest that stroke survivors should receive medical clearance from a medical professional before participating in an exercise programme, which is followed by most ERS through the referral pathways, although some schemes allow self-referrals. This contrasted with schemes in Scotland, where only half required a referral from a health professional.^18^ The recommended frequency of sessions is between three to five per week.^34^ However, 79% of responders reported session frequencies outside of these recommendations. For ERS offering less than three sessions per week, stroke survivors may be able to receive more benefit from a greater dose of exercise, and schemes offering more than five sessions per week may be wasting resources. However, most ERS followed the recommended session duration of 60 minutes and provided a wide range of exercise types for participants. Although some stroke-specific exercise recommendations are being met by current ERS, changes must be made to ensure stroke survivors are receiving the recommended dose of exercise.

Fitness instructors have a central role in ERS and are often the sole providers of exercise within these schemes. All programmes required fitness instructors to have at least a level three qualification, which is aligned to current ERS guidance.^11^ Stroke-specific exercise guidelines recommend that fitness instructors with appropriate qualifications should provide exercise for stroke survivors, but links to stroke clinicians must be available.^34^ In many ERS, stroke clinicians were not included, meaning stroke expertise is not readily available. Previous research into ERS has highlighted potential safety concerns with fitness instructors leading exercise sessions, due to their lack of stroke expertise.^15^ Although some stroke-specific qualifications are available to fitness instructors,^35^ these are not available throughout the UK. In Scotland, none of the fitness instructors involved with stroke-specific exercise programmes held a formal stroke qualification.^18^ Given the lack of nationwide training, ERS should adopt a collaborative approach with health services to utilise the stroke expertise of clinicians. Research in other countries have found that including health professionals in community exercise programmes is viewed positively by stroke survivors.^36^ Fitness instructors should continue to have a central role in ERS for stroke survivors; however, stroke-specific training should be made more widely available and links to healthcare services should be established.

The findings from this study could explain the low utilisation of ERS by stroke survivors in the UK. Data from the National ReferAll database showed that only 0.1% of ERS referrals were for stroke-specific schemes.^17^ First of all, the number of stroke-specific ERS is relatively low, with only 17 of the 147 responding local authorities providing this service. In general ERS many stroke survivors will be ineligible due to the lasting effects of the stroke. Even for stroke-specific schemes, it is unclear if moderately disabled stroke survivors are eligible, or if these or designed for those with mild disabilities. The barriers to stroke survivors accessing community exercise services have long been established,^30^ but ERS have not been designed to overcome these. Assistance with transport was provided in only three of the responding ERS, and the financial cost to participate may be unfeasible for some stroke survivors in many local authorities. If a stroke survivor can overcome these barriers, the ERS may not be able to provide enough support due to high service user to supervisor ratios, and lack of staff with stroke expertise, which may result in poor adherence. Significant changes to the design and delivery of ERS must be made to improve the accessibility to stroke survivors.

### Limitations

Due to the lack of a central database of ERS in the UK the number of schemes in England is unknown, making the true coverage of this study unclear. This problem has also affected similar studies.^13,26^ Furthermore, the lack of a central database meant that recruitment materials had to be sent to the local authority who then forwarded materials to the specific ERS team, which had a negative effect on response rate, with around half of local authorities engaging with the survey. It is possible that the characteristics of responding versus non-responding local authorities are different, thus response bias may be present. Another limitation is the reliance on self-reported data. Other studies have evaluated ERS using online materials.^14^ However, much of the data collected through this survey (i.e., capacity, funding, etc) would not be publicly available, making it very difficult to validate. Finally, the exclusion of Scotland, Wales, and Northern Ireland means the current accessibility in these countries is unknown. It should not be assumed that accessibility in England represents other nations within the UK.

### Implications for practice, policy, and future research

There is a need to establish a central database of ERS in the UK to allow better evaluation of these services. Until this is done, research in this area will continue to produce partial coverage. There is an urgent need to standardise the delivery of ERS. This study has identified wide variation in the design and delivery of ERS in England, which may be perpetuating health inequalities. Exercise is now recommended by clinical guidelines, and all stroke survivors should have equal access to a standardised programme of exercise. This study has shown that stroke-specific services already exist, but these need to be expanded throughout the UK. To facilitate this expansion, availability of stroke-specific training for fitness instructor needs to be more widely available. Two of the main barriers to stroke survivors exercising in the community are financial cost and lack of transport,^30^ which are not being negated by current ERS provision. Services should prioritise securing funding to provide transport for those who need it and reduce costs of participation to a minimum.

Although previous research has investigated stroke survivors experiences of participating in an ERS, the level of disability of participants was not reported.^15,16^ Future research should aim to evaluate the level of disability of stroke survivors who take part in ERS, and explore if experiences differ across different levels of disability. Research should also aim to investigate if socioeconomic status influences a stroke survivors’ likelihood of engaging with an ERS. There has yet to be a study investigating the clinical efficacy of ERS for stroke survivors in the UK. Future studies should aim to investigate the effects on a wide range of clinical outcomes, including cognitive and psychosocial.

## Conclusion

ERS are widely available across England, and the majority are open to stroke survivors. However, stroke-specific services are very limited. Significant variations were identified in the design and delivery of ERS, the only two consistencies found between schemes was the involvement of fitness instructors and lack of transport provision. Known barriers to stroke survivors exercising in the community are present in the majority of ERS in England. All these factors are contributing to the under-utilisation of ERS by stroke survivors. Standardisation of stroke-specific ERS across the UK would result in more equitable access to services and likely increased utilisation.

## List of abbreviations

ERS: Exercise referral scheme
UK: United Kingdom
NICE: National Institute for Health and Care Excellence

## Declarations

### Ethics approval and consent to participate

Ethical approval was granted by the Lancaster University Faculty of Health and Medicine Research Ethics Committee (FHM-2024-4432-RECR-2). All participants provided informed consent to participate in the study. The study was conducted in accordance with the Declaration of Helsinki.

### Consent for publication

Not applicable

### Availability of data and materials

All data generated by this study is publicly available from: https://osf.io/yxkew/overview

### Competing interests

The authors declare that they have no competing interests

## Funding

The primary author (JS) is supported by a PhD studentship provided by Lancaster University Faculty of Health and Medicine. The funder had no other role in the study.

## Authors contributions

Conceptualisation - JS

Methodology - JS

Software - JS

Formal analysis - JS

Investigation - JS

Data curation - JS

Writing (original draft) - JS

Writing (review & editing) - PN, HJ

Visualisation - JS

Supervision - PN, DT, CH, HJ

## Data Availability

All data produced in the present study are available upon reasonable request to the authors.

## Acknowledgements

Not applicable

## Notes

### Competing Interest Statement

The authors have declared no competing interest.

### Author Declarations

Faculty of Health and Medicine Research Ethics Committee of Lancaster University gave ethical approval for this work (FHM-2024-4432-RECR-2).

